# Cohort profile: CROSS-TRACKS – a population-based open cohort across healthcare sectors in Denmark

**DOI:** 10.1101/2020.05.13.20100263

**Authors:** Anders Hammerich Riis, Pia Kjær Kristensen, Matilde Grøndahl Petersen, Ninna Hinchely Ebdrup, Simon Meyer Lauritsen, Marianne Johansson Jørgensen

## Abstract

**Purpose:** This paper describes the open cohort CROSS-TRACKS, which was set up in Denmark in 2015. The cohort comprises population-based data from primary care, secondary care, and national registries. In addition, the paper outlines the new research opportunities provided by the cohort; the possibility to study patient pathways and transitions across sectors while adjusting for sociodemographic characteristics. The new data source is especially interesting for studies of potentially preventable admissions and readmissions.

**Participants:** A total of 221,283 individuals resided in the four Danish municipalities that constituted the catchment area of Horsens Regional Hospital in 2012–2018. A total of 96% of the population used primary care, 35% received at least one transfer payment, and 66% was in contact with a hospital at least once in the period. Additional clinical information is available for hospital contacts (eg alcohol intake, smoking status, body mass index, and blood pressure). A total of 27% (n=9,986) of individuals aged ≥65 years had at least one potentially preventable hospital admission, and 69% (n=6,881) of these individuals had more than one.

**Findings to date:** The cohort is currently used for research projects in epidemiology and artificial intelligence. These projects comprise a prediction model for potentially preventable hospital admissions, a clinical decision support system based on artificial intelligence, prevention of medication errors in the transition between sectors, health behaviour and sociodemographic characteristics of men and women prior to fertility treatment, and a recently published study applying machine learning methods for early detection of sepsis.

**Future plans:** The CROSS-TRACKS cohort will be expanded to comprise the entire Central Denmark Region consisting of 1.3 million residents. The cohort can provide new knowledge on how to best organise interventions across healthcare sectors and prevent potentially preventable hospital admissions. Such knowledge would benefit both the individual citizen and society as a whole.

**Strengths and limitations of this study:**

1. The CROSS-TRACKS cohort is a population-based open cohort containing routinely collected data from both primary and secondary care combined with sociodemographic register data.
2. The cohort is readily available for research projects: the data sources have already been linked, patient pathways have been identified, and data have been remodelled and prepared for cutting-edge use of artificial intelligence and to enable easier and simpler querying across source systems.
3. The cohort is currently limited to the catchment area of Horsens Regional Hospital, but it covers both urban and rural areas and it has a total population of 221,283 individuals by 2018.
4. The cohort will be expanded to include additional catchment areas in the Central Denmark Region, comprising a total population of 1.3 million individuals.
5. The cohort offers new opportunities for health research by tracking the patients’ pathway across healthcare sectors to provide new knowledge on the best way to organise future interventions.

## Introduction

An increasingly ageing population and limited resources place new demands on society and health care[1]. The global population of individuals aged 65+ years is projected to increase from 8.2% in 2015 to 15.9% in 2050, corresponding to > 1.5 billion people[2]. The elderly population has higher needs for health care compared to younger citizens[3,4]. For example, the number of hospitals admissions have increased by 45% from 2006 to 2018 among Danes aged 65+ years[5]. Therefore, Denmark and many other countries have reorganised the healthcare systems; they have reduced the number of hospital beds and shortened the length of hospital stays. The total number of days in a hospital bed has decreased by 16% from 2006 to 2018[5]. Therefore, an increasing part of the healthcare, treatment, and rehabilitation services are provided by general practitioners (GP) and the municipalities in the primary care setting.

One focus has been directed towards reducing the number of potentially avoidable readmissions[6]. In the US, almost 20% of discharged patients are readmitted to hospital within 30 days[7]. This is costly to the individual citizens and to society. Special attention has been turned towards potentially preventable admissions and readmissions. These are often seen for ambulatory care sensitive conditions (ACSCs), which comprise conditions that can usually be treated and prevented in the primary care setting[8]. More than 20% of hospital admissions among elderly US residents in nursing homes are estimated to concern ACSCs[9]. In Denmark, 15% of all hospitalisations of the elderly population are expected to be preventable[10]. The rates of potentially preventable hospital admissions are high in Denmark compared to other European countries[11]. Furthermore, a recent study found higher rates of potentially preventable hospitalisations and readmissions for chronic medical conditions in Denmark compared to the US[12]. Consequently, there seems to be room for improvement in our approach to preventable admissions and readmissions.

Several interventions have aimed to reduce the risk of readmissions[13] and to improve the care for discharged patients in the primary care setting[14]. Most of these focused on the pre-discharge period (eg patient education and discharge planning) or the post-discharge period (eg follow-up telephone calls and home visits), whereas only few interventions bridged over both sectors (eg patient-centred discharge instructions and transition coaching). The interventions bridging across sectors were most effective[13]. This calls for studies of patient transitions across sectors in a population-based setting.

CROSS-TRACKS[15] is an open cohort comprising routinely collected administrative data from primary care, secondary care, and national registries. The combined data sources allow us to explore key factors such as sociodemographic characteristics, labour market affiliation, social transfers, home-based nurse care, GP contacts, and potential links between these factors and the use of services in secondary care.

The cohort is a valuable tool for studying patient pathways across sectors, including the patterns that characterise potentially preventable hospital admissions. This paper describes the new opportunities in healthcare research that are emerging from the comprehensive CROSS-TRACKS cohort.

## Cohort description

### Setting

The tax-funded healthcare system in Denmark provides free access for all citizens to healthcare services. The five Danish regions are the backbone of the healthcare system, and they are responsible for organising and operating the somatic and psychiatric hospitals (Figure 1). Furthermore, the regions reimburse GPs and specialists in private practice (eg physiotherapists, psychologists, or dentists) for dispensed prescriptions and services provided[16]. The GPs can refer citizens to a wide range of welfare services (eg home care and rehabilitation) that are provided in the primary care setting by the 98 municipalities under the regions[16]. The catchment area usually comprises the municipalities in the geographical area around the hospital.

**Figure 1.**
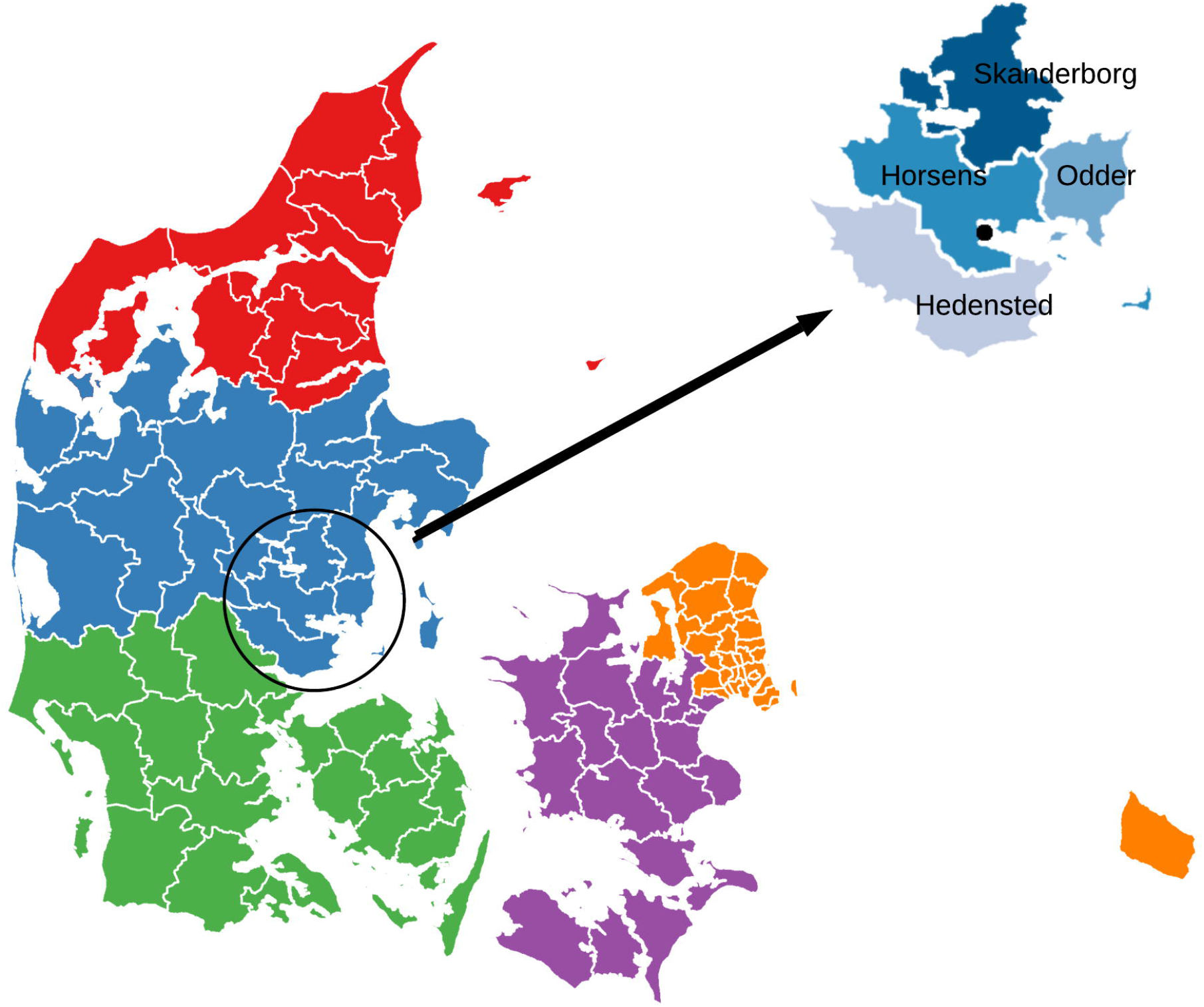
The five Danish regions and the 98 municipalities. The black circle indicates the four municipalities (Hedensted, Horsens, Odder and Skanderborg) included in the CROSS-TRACKS cohort, and the black dot indicates the location of Horsens Regional Hospital.

The GP is the primary entry point into the healthcare system for Danish citizens, and the GP acts as a gatekeeper to secondary care[17], but the citizens can contact the prehospital directly for emergencies. Citizens have unrestricted access to dental care, which is free of charge for citizens below the age of 18. After hospital discharge, patients may be referred to the GP or a triage manager in the municipality for further treatment, care, or rehabilitation.

All Danish citizens receive a unique identifier at birth or immigration. This number is also referred to as the civil registration (CPR) number[18]. The CPR number enables electronic linkage for all data in the cohort across registries and databases. The cohort data is securely stored at the data warehouse of the Central Denmark Region in accordance with data protection regulations. The cohort is readily available for research projects as the data sources have already been linked and patient pathways have been identified. Further, the data have been remodelled according to the dimensional modelling principles laid out by Kimball and Ross[19], to provide a simple and consistent query interface across source systems.

## Study population

The CROSS-TRACKS cohort consists of all citizens aged 18+ years residing in the catchment area of Horsens Regional Hospital, which comprises Hedensted, Horsens, Odder and Skanderborg municipalities (Figure 1). The inclusion period started on 1 September 2012 (cohort entry date) and will last through 2022 (Figure 2). Individuals are included on their 18th birthday or when moving to one of the four included municipalities. Individuals are followed from the date of inclusion until five years after the end of the inclusion period (31 December 2027) or date of death. Individuals moving away from these municipalities are followed for five years after the date of moving.

**Figure 2.**
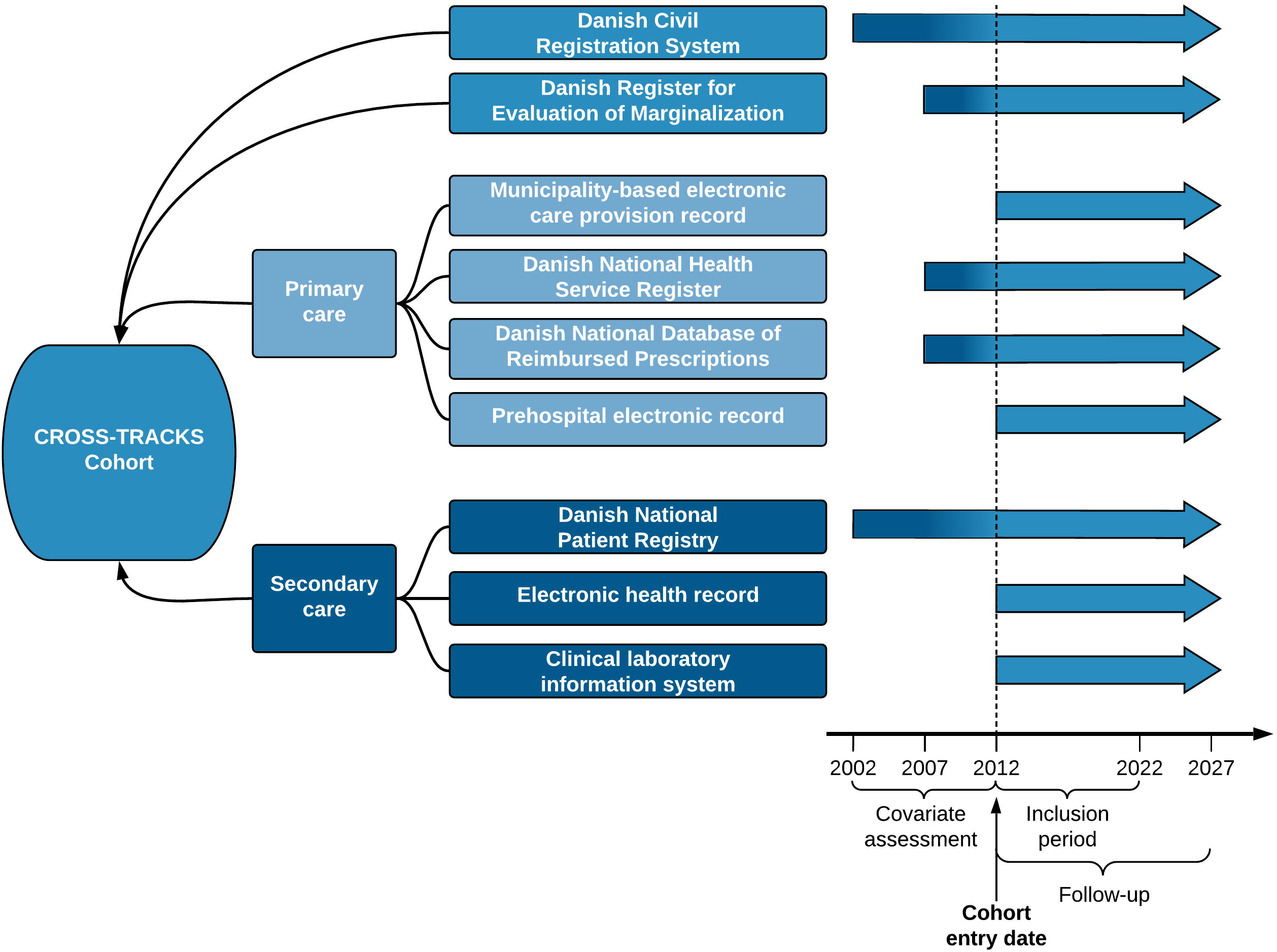
Data availability for the CROSS-TRACKS cohort in Denmark.

Patient characteristics are time-dependent because disease develops over time, and this affects the number and frequency of contacts with the healthcare system. The use of nationwide data sources ensures optimal adjustment for potential differences in patient characteristics, even for individuals living outside the included municipalities during part of the follow-up period. Moreover, we use look-back periods to assess covariates prior to cohort entry date[20]. The length of look-back periods can be found in Figure 2.

## Data collection

Individuals are included in the cohort according to the inclusion criteria shown in the flow chart in Figure 3.

**Figure 3.**
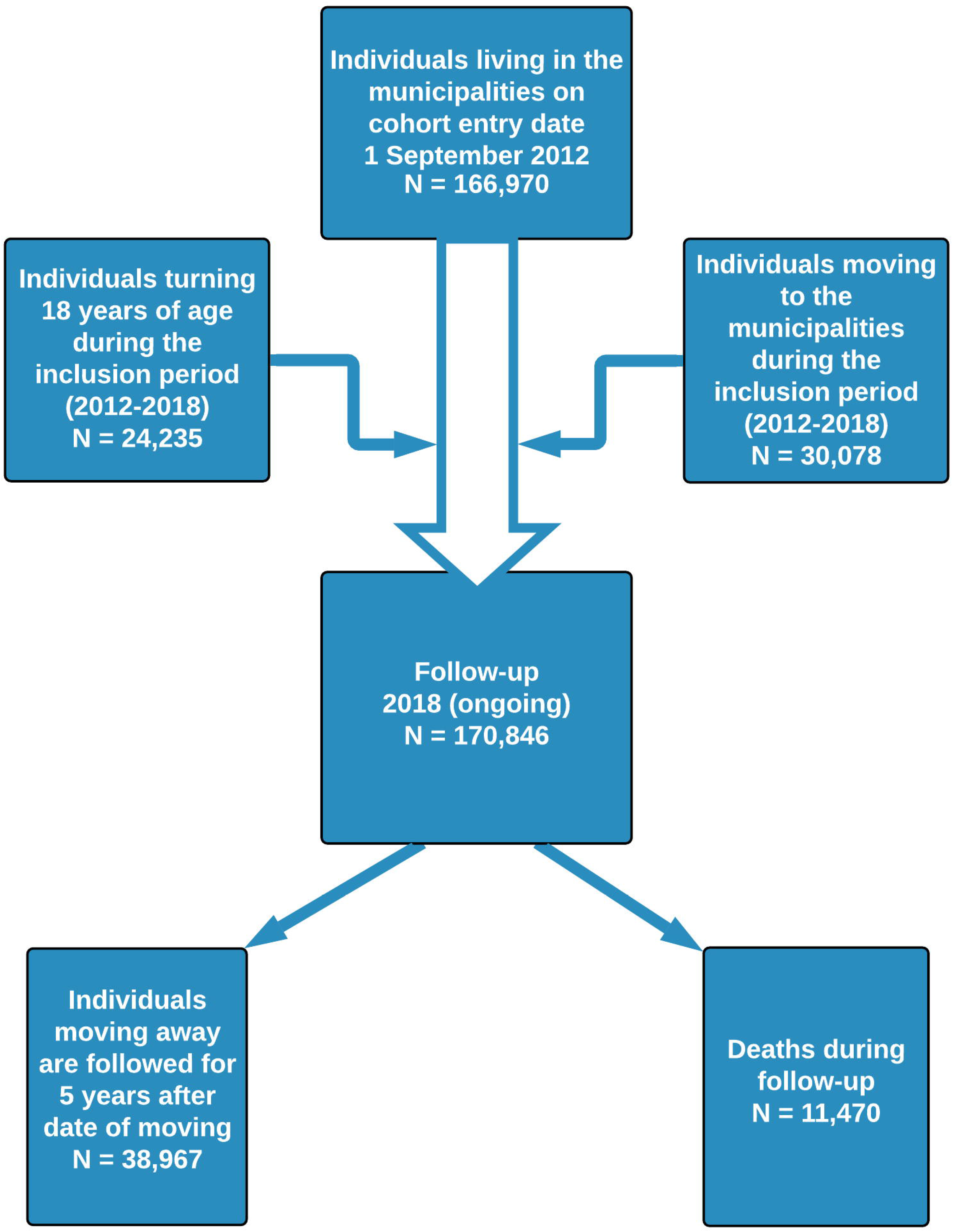
Flow chart of individuals included in the CROSS-TRACKS cohort in Denmark 2012–2018.

### Administrative and health registries

The CPR number originates from the Danish Civil Registration System (CRS)[18], which also provides date of birth, gender, vital status, civil status, migration, and place of residence. The CRS is updated on a daily basis with information on vital status and place of residence; this ensures complete follow-up for all individuals. The CROSS-TRACKS cohort includes data from the CRS from 1 September 2002, which enables a 10-year look-back period prior to each individual’s date of inclusion.

Weekly recordings of social benefits and other transfer incomes are retrieved from the Danish Register for Evaluation of Marginalization (DREAM)[21]. DREAM can be used to categorise individuals into unemployed, on sick leave, receiving part-time benefit, receiving disability pension, or receiving retirement pension[22].

#### Primary care

The municipality-based electronic care provision record (ECPR) holds data on the health services granted to the citizens. These services comprise municipality-based practical help, rehabilitation assistance, personal care, and home visits by a community nurse. The data includes the period (start and end dates) and total amount of time assigned to the services.

The Danish National Health Service Register (DNHSR) holds information on contacts with healthcare professionals (eg GP, dentist), type of consultation (eg ordinary consultations, home visits, telephone consultations, electronic consultations), services provided, and laboratory tests performed[23].

The Danish National Database of Reimbursed Prescriptions (DNDRP) contains detailed information on prescribed drugs that are partially reimbursed by the regions. This includes information on date of the redemption, number of pills, dose, number of defined daily doses, name and type of drug according to the Anatomical Therapeutic Chemical classification system[24].

The prehospital electronic record holds information on treatment and transportation of patients by ambulance, medical cars, or helicopters. Registrations include urgency level, date and time of departure and arrival, treatments, medications, and a free-text field for additional information that may help ensure optimal transfer of patients to the hospital[25].

#### Secondary care

The Danish National Patient Registry (DNPR) encompasses all hospital contacts, including admission date, discharge date, and outpatient visits. Diagnoses, surgical procedures, and examinations are registered according to the International Classification of Diseases[26]. The main reason for each hospital contact is registered as the primary diagnosis, whereas secondary diagnoses are stated as other diseases in relation to the contact. Diagnoses registered in the DNPR can be used to identify individuals admitted to hospital with a primary ACSC-related diagnosis. In addition, the Charlson Comorbidity Index (CCI) score is calculated based on 19 conditions[27].

The electronic health record (EHR) in the Central Denmark Region consists of healthcare-related registrations linked with date and time. The DNPR is based on registrations in the EHR, but this record contains additional data that are not transferred to the DNPR. This includes registrations on triage, patient’s pulse, blood pressure, respiration frequency, and microbiology test results. Some of these are used for early detection of critical illness. The EHR can also be used to identify patient-related lifestyle factors, such as diet, smoking and alcohol habits, and physical activity. Furthermore, hospital-based drug treatment is registered in the EHR by the doctors prescribing the drug and the nurses administering the drug by dispensing the medicine to the patients.

The clinical laboratory information system (LABKA) consists of test results from blood and urine samples drawn by health professionals (in both primary care and hospitals) and analysed in clinical laboratories[28]. Registrations include date of sample, health professional or hospital department requesting the analysis, requested assay or biomarker, and corresponding Nomenclature, Properties and Units (NPU) code.

An overview of data sources and availability can be seen in Figure 2 and Table 2. The codes used to extract the characteristics of citizens can be found in Table 1 in the supplementary material.

**Table 1.**
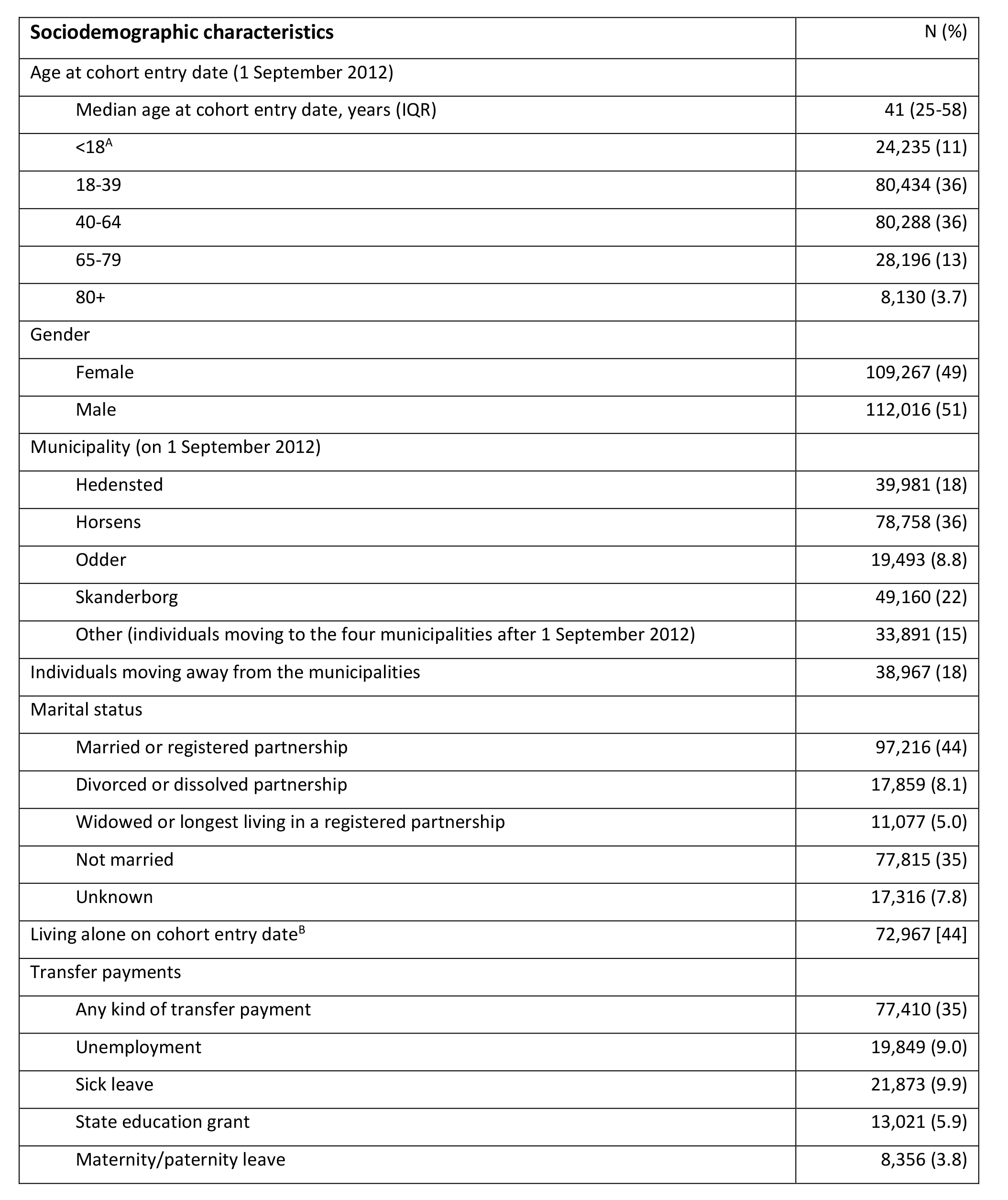
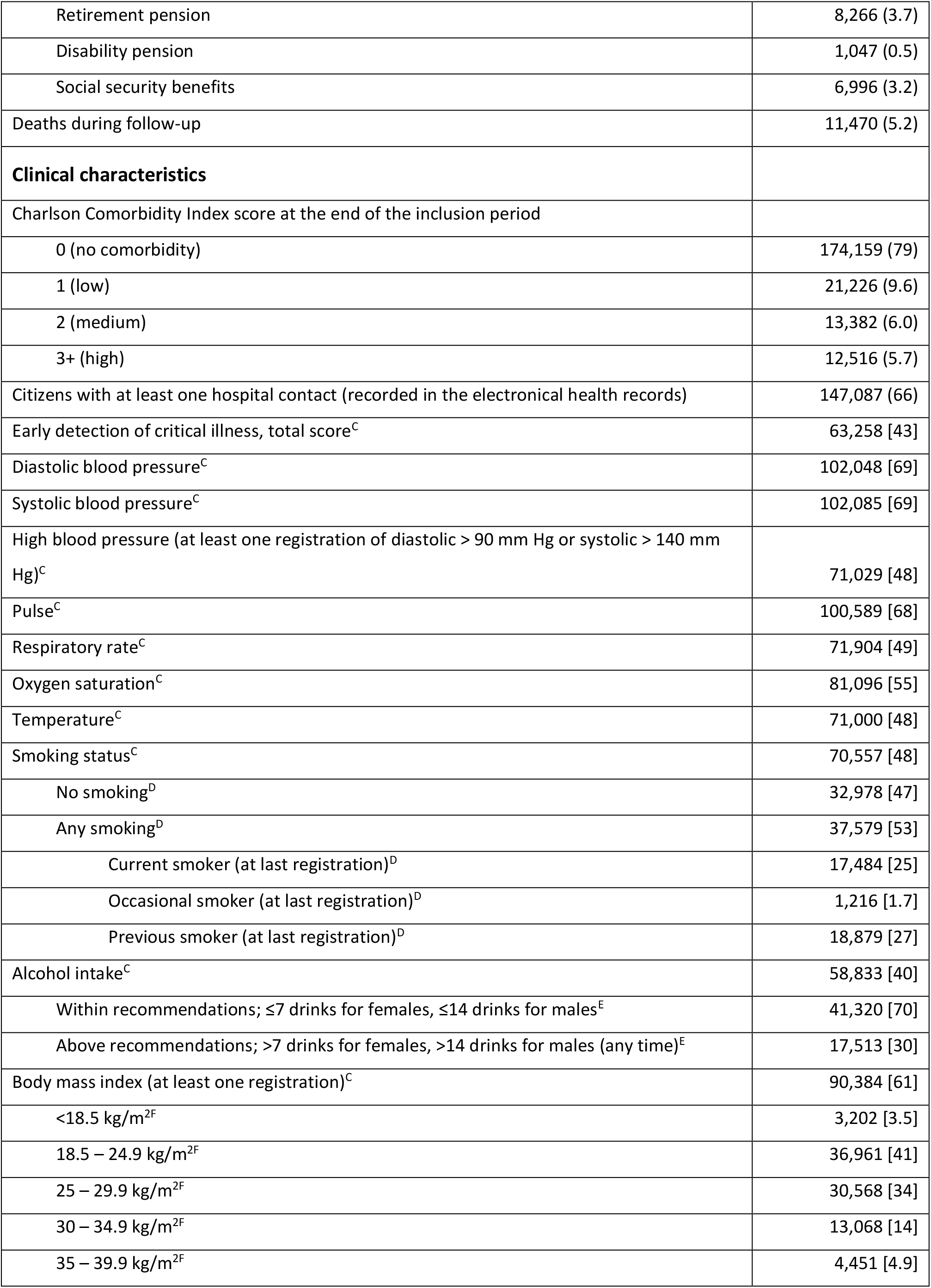
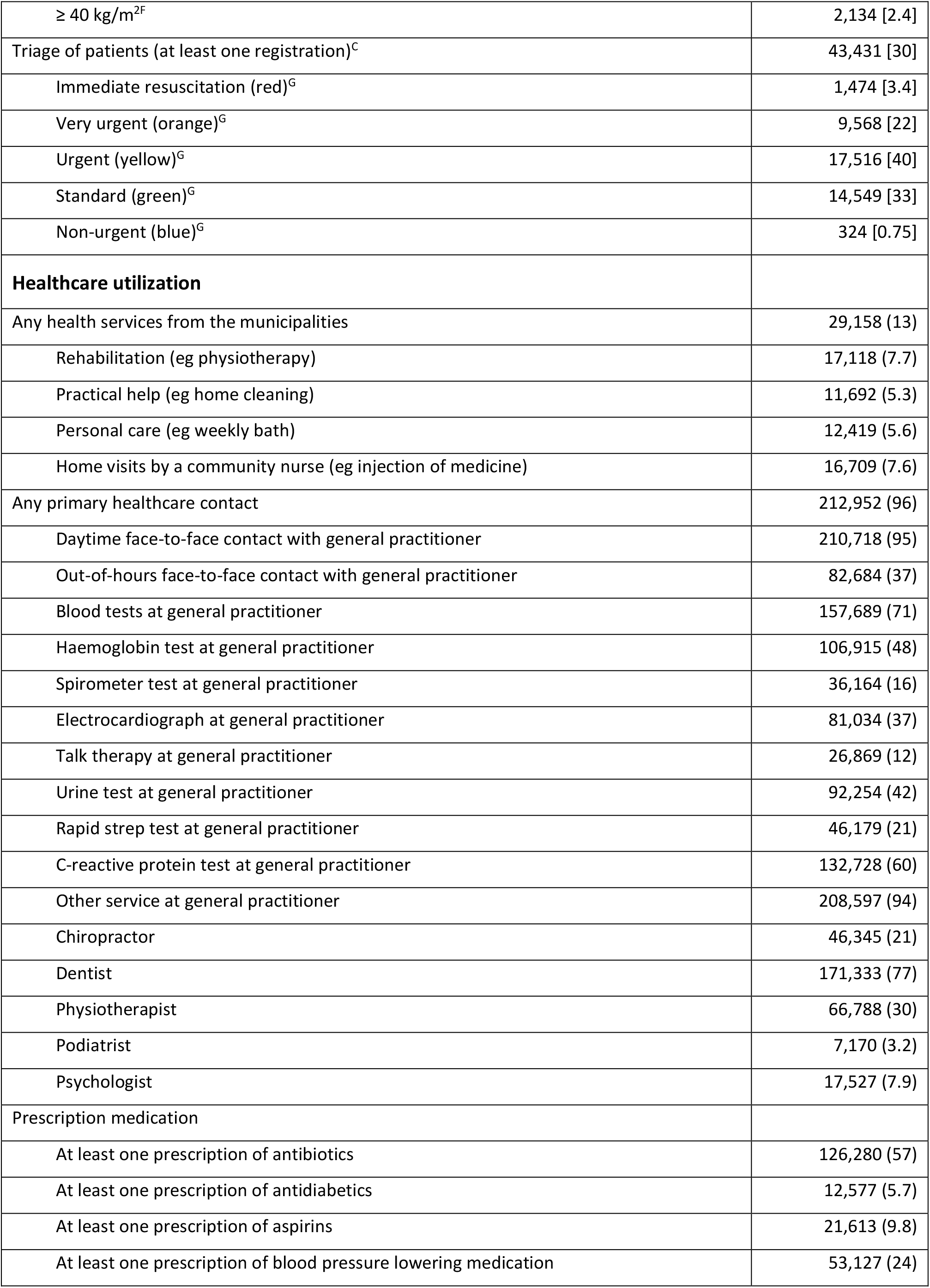
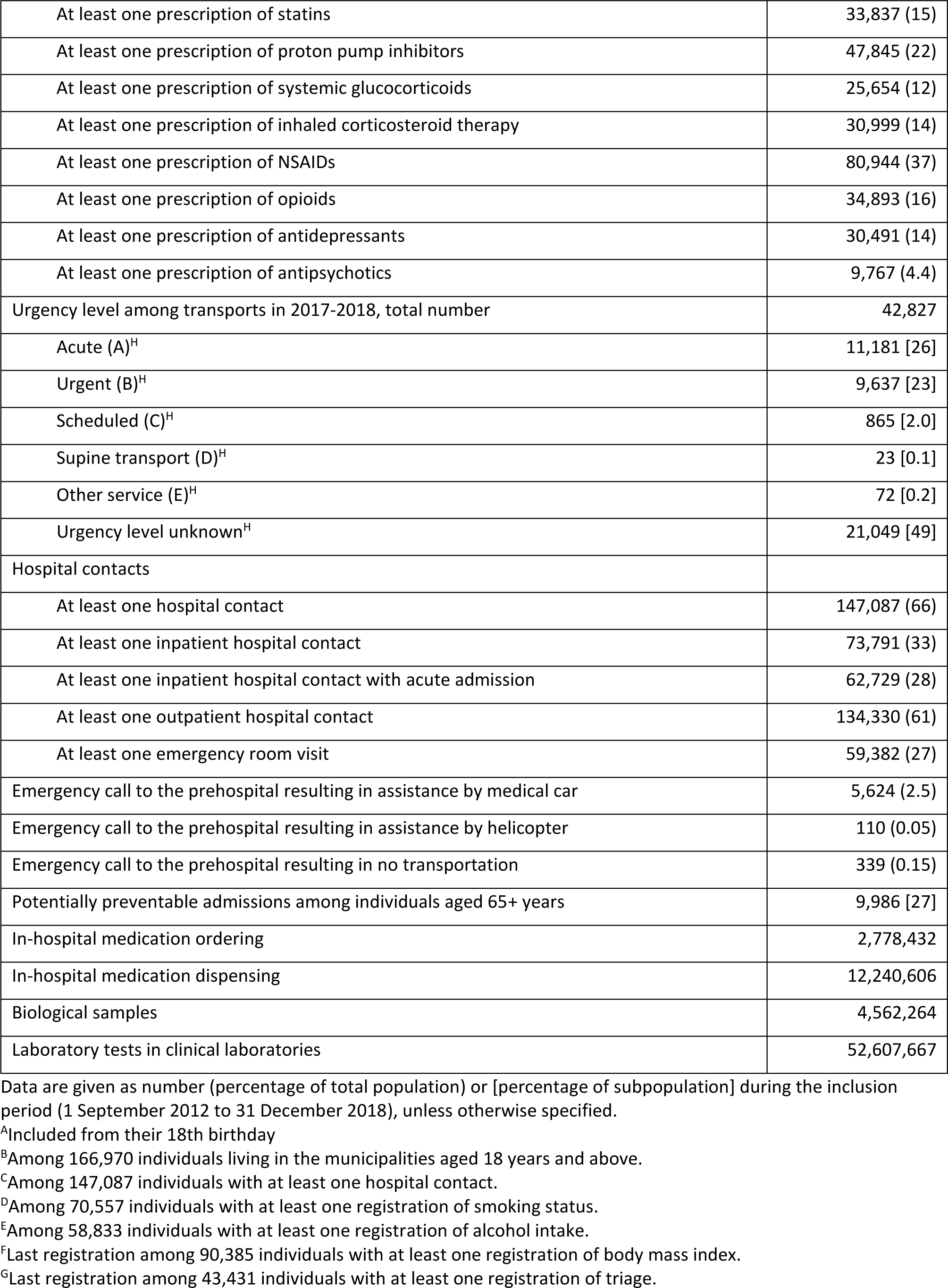
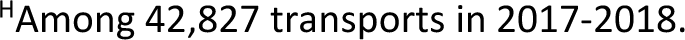
Sociodemographic characteristics, clinical characteristics, and healthcare utilization among citizens in the four included Danish municipalities (Hedensted, Horsens, Odder and Skanderborg) in 2012–2018

Although the EHR data in the data warehouse of the Central Denmark Region is updated on a daily basis, the combined cohort consisting of all data sources is only updated annually. The cohort is dynamic in the sense that the data quality is continuously improved as new research projects build on new sections of the cohort. See Table 2 in the supplementary data for a description on how registrations on body mass index (BMI) was cleaned before use.

**Table 2.**
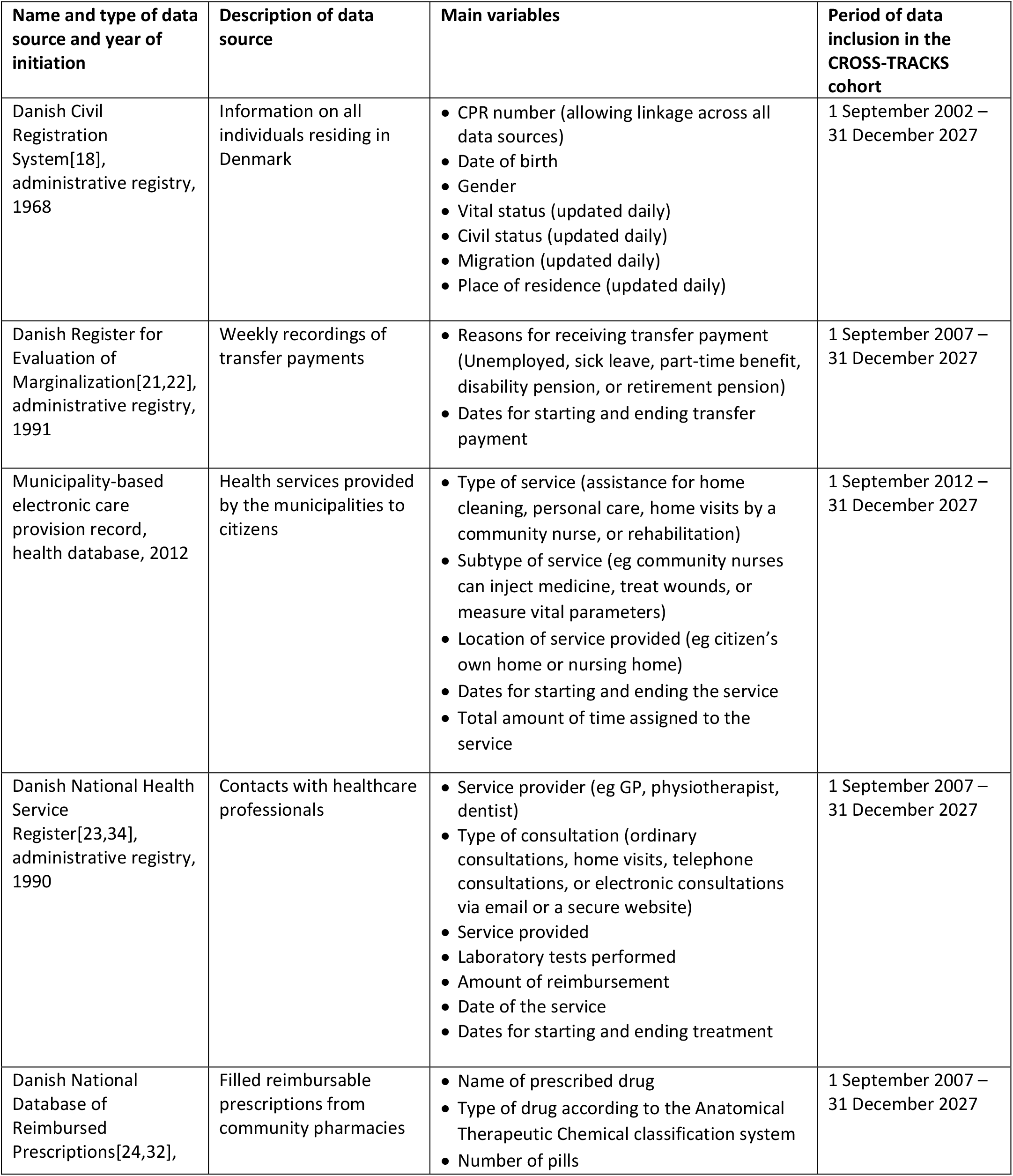
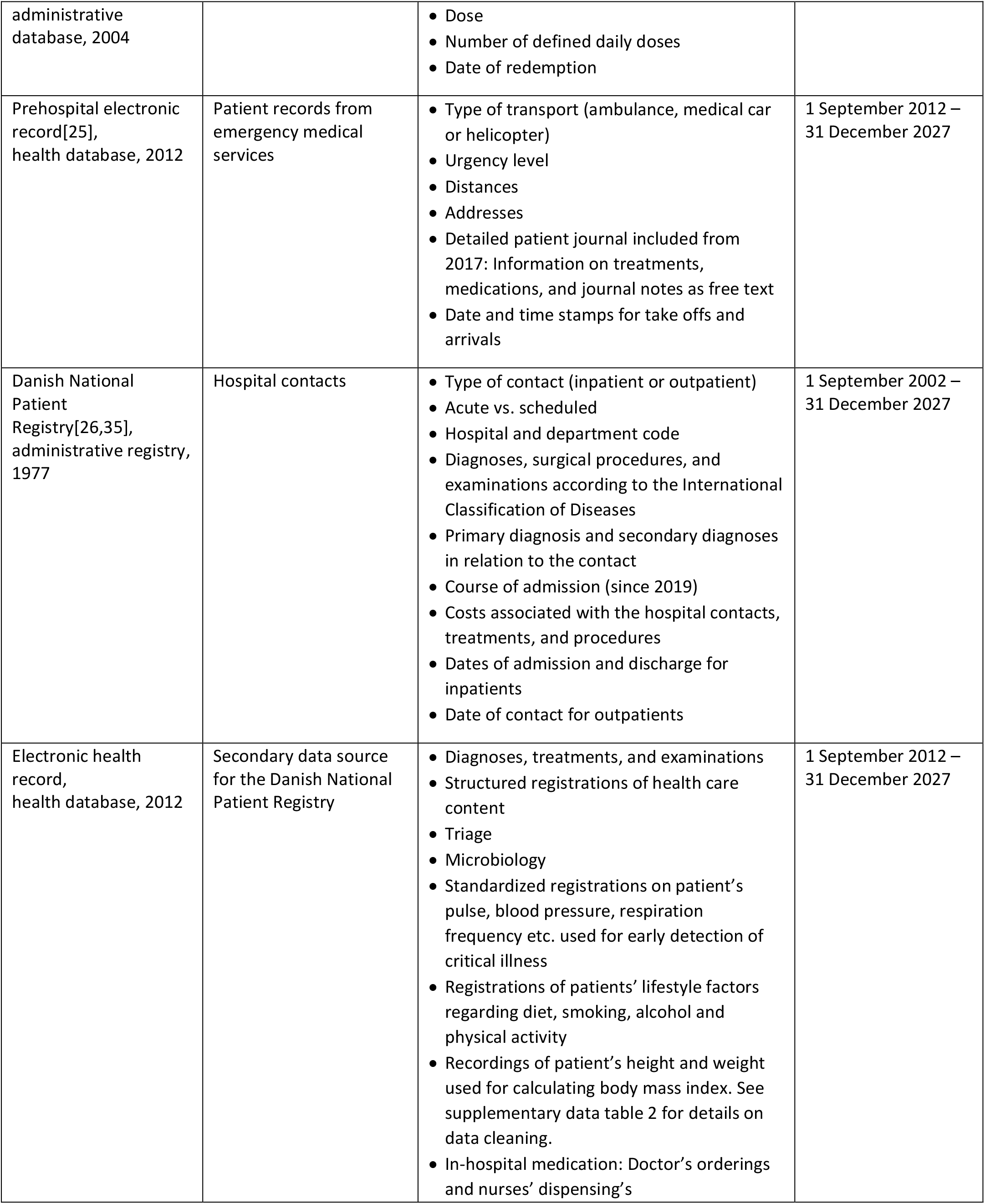
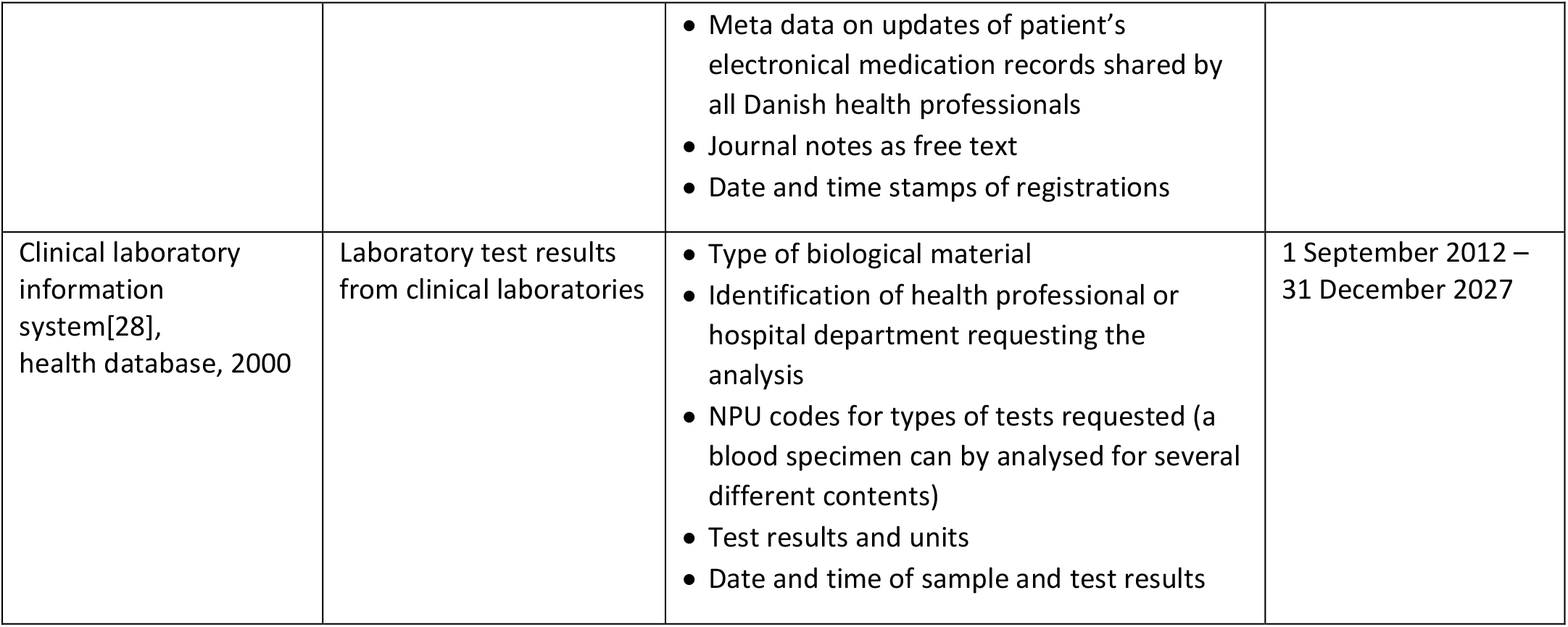
Administrative and health registries included in the CROSS-TRACKS cohort[18], [21,22] [23,34] [24,32] [25] [26,35] [28]

### Population characteristics

On the cohort entry date, 166,970 individuals aged 18+ years resided in the four municipalities. From 1 September 2012 through 2018, 24,235 individuals turned 18 years of age, and 30,078 moved to one of the municipalities. This yields a total population of 221,283 individuals in the open cohort (Figure 3). The main characteristics of citizens in the cohort are shown in Table 1. Individuals aged below 18 years appear in the table, but they are not included in the cohort before their 18th birthday.

The catchment area of Horsens Regional hospital contained one health centre (located in Skanderborg), 69 GPs, 20 specialists in private practice, 11 chiropractors, 52 dentists, 45 physiotherapists, 25 podiatrists, and 25 psychologists.

The largest municipality was Horsens with 78,758 citizens, corresponding to 36% of the population on 1 September 2012. Skanderborg had 49,160 (22%) citizens, Hedensted had 39,981 (18%) citizens, and Odder had 19,493 (9%) citizens. In the beginning of the inclusion period, Horsens was the only urban municipality, but Skanderborg went from rural to urban during the period.

The median age on cohort entry date was 41 years (IQR: 25–58 years) with an equal distribution of gender. The proportion of citizens who were married or living in a registered partnership was 44%, and 44% of the citizens aged 18 years and above were living alone on cohort entry date.

During the inclusion period from 1 September 2012 through 2018, 35% of the total population received some kind of transfer payment. Sick leave was the predominant reason with 10%, which was followed by unemployment among 9% of the population.

A total of 21% had a CCI score of 1 or above at the end of the inclusion period. For the part of the population (66%) with a hospital contact in the period from 1 September 2012 through 2018, we used registrations in the EHR to obtain the clinical characteristics. Early detection of critical illness was registered in the EHR among 43% of the population with at least one hospital contact. Registrations of blood pressure were used to identify patients with at least one registration of high blood pressure (49%). Smoking status was recorded in 48% of individuals with at least one hospital contact, and 53% of these reported any smoking. Alcohol intake was registered in 40% with at least one hospital contact, and 30% of these had at least one incidence of intake above the recommendations of the health authorities, which is up to 7 drinks per week for females and 14 drinks per week for males[29]. Of the 90,385 patients with information on BMI, 50,222 (56%) were overweight (BMI ≥25 kg/m^2^).

### Healthcare utilisation

Almost everyone in the population received at least one service provided from primary care during the inclusion period. Daytime GP services were utilised by 95% of the population, dental services by 77%, and psychologist services by 8% during the inclusion period. Municipality-based care was provided to 13% of the citizens receiving rehabilitation assistance (8%), and nursing care (8%) was the most frequent service.

Antibiotics was the most frequently used type of prescription medication; 57% of the population redeemed at least one antibiotics prescription during the inclusion period. NSAID prescriptions was redeemed by 37%, and blood pressure lowering medication was used by 24% of the population.

A total of 42,827 patient transportations were made by the prehospital in the period from 2017 to 2018; 26% were acute, 23% were urgent, and 49% had unknown urgency level. The transports concerned 18,671 unique individuals, and 7,786 individuals had more than one transport. Overall 66% of the population had at least one hospital contact during the inclusion period, 33% had an inpatient contact, 61% had an outpatient visit, and 27% had an emergency room visit.

In the population of individuals aged 65+ years, 9,986 (27%) had at least one potentially preventable hospital admission during the inclusion period; 6,881 (69%) of these had more than one potentially preventable hospitalization.

Almost three million medications were ordered, and about 12 million medications were dispensed in the hospitals. Overall, 4.5 million biological samples were drawn, and 52 million laboratory tests were performed on these samples.

## Findings to date

The CROSS-TRACKS cohort was established to gain greater research-based knowledge on the causes of potentially preventable (re)admissions by tracking patient pathways across healthcare sectors. The new knowledge can be used to strengthen the collaboration across sectors and to target joint efforts.

The cohort is currently being used for several ongoing health research projects. These include a prediction model for potentially preventable hospital admissions, a clinical decision support system based on artificial intelligence, prevention of medication errors in the transition between sectors, and health behaviour and sociodemographic characteristics of men and women in fertility treatment. Another study has applied machine learning methods for early detection of sepsis and has identified potential for initiating interventions earlier[30]. In continuation of this study, the authors used explainable artificial intelligence to develop an early-warning score system for detection of acute critical illness[31]. This study adds transparency in artificial intelligence by explaining which relevant electronic health records data predicts critical illness.

## Strengths and limitations

CROSS-TRACKS is the first cohort to provide readily available data from primary and secondary care for research projects: the data sources have already been linked, and patient pathways have been identified. A major strength of the cohort is the completeness and the high quality of the available data, which is obtained through the prospective collection of population-based data. An additional strength is that the unique Danish CPR number allows linkage across registers and complete follow-up for all individuals; this offers the possibility to monitor disease progression over time. The clinical information about the patients can be followed through the EHR when they live in the Central Denmark Region. If they move away, the DNPR can be used to follow the individual’s disease development. Additionally, the 10-year look-back period and the follow-up through 2027 hold the possibility of following citizens for up to 25 years.

Given the detailed information on medication in both sectors, this cohort presents the complete history of registry-based drug use for each individual; this fosters rich opportunities for adjusting for confounders. Furthermore, the combination of psychiatric inpatient contacts, antidepressants and antipsychotics prescriptions, and talk therapy by GPs and psychologists offers potential for identifying psychiatric disease. In addition, several validation studies have reported high validity of the Danish registry data[21,26,32]. The cohort itself forms a basis for easy validation as the free-text registrations in the EHR are usually regarded as the gold standard to which registry information is compared.

Nevertheless, we note some limitations of the cohort. Not all data sources have yet been validated, and there are missing data on lifestyle characteristics, as they may not have been reported for all citizens in contact with a hospital. However, investigator bias is limited since much of the data is recorded by clinicians during diagnosis and treatment, and not for research purposes. The information on medication use in primary care is based on reimbursable redeemed prescriptions, whereas no information on over-the-counter medication is available. However, citizens with regular medication use have economic incentives to acquire the prescribed drugs because of the reimbursement option. Furthermore, a study on the use of aspirin and NSAIDs found that the potential for identifying medication use through prescription registries in Denmark is high[33].

Another limitation is the missing data on treatments by private psychologists, physiotherapists, and chiropractors; these services are not covered by the healthcare system. However, the private healthcare sector is small in Denmark and is mostly based on services eligible for reimbursement by the regions, and these services are thereby included in the registries. Additionally, private hospitals report to the DNPR[16], which also contributes to maintaining the high completeness of registry data.

The municipalities use different suppliers for reporting to the ECPR. This means that it requires extensive data management to streamline the information, and this could be a potential limitation. The inclusion of ECPR data is conducted in close collaboration with the municipalities and their data managers to ensure correct interpretation and high validity of the data.

Only the catchment area for Horsens Regional hospital is included in the cohort, and this may compromise the generalisability. However, the catchment area includes both urban and rural municipalities. In addition, citizen contacts with Aarhus University Hospital, which is located in the Central Denmark Region, are also included in the cohort through the recordings in the EHR. Because of the structure of the Danish national registries and databases, the cohort can easily be extended to comprise additional municipalities and regions and ultimately all of Denmark. Starting in 2020, we will expand the cohort to include all 1.3 million residents in the Central Denmark Region. This region consists of one university hospital, one psychiatric hospital, four hospitals with 24-hour emergency department, four hospitals without 24-hour emergency department, and ten health centres and acute clinics[16].

The CROSS-TRACKS cohort provides valuable new opportunities for health research across primary and secondary care in the future; this is expected to benefit both the individual citizens and society as a whole. The cohort can contribute with new knowledge on how to organise interventions across sectors to reduce the number of potentially preventable (re)admissions.

## Collaboration

Access to CROSS-TRACKS data can be granted for research and analysis projects if certain criteria are met, and an application has been submitted to the steering committee. The committee strongly encourages national and international collaboration. For more information on access criteria and application form, see the website www.tvaerspor.dk (currently in Danish).

## Further details

### Contributors

AHR, PKK, and MJJ wrote the first draft of the manuscript. MGP was responsible for data access. AHR performed the statistical analyses. All authors provided critical review of the manuscript and approved the final version.

### Funding

The project was supported by Horsens Regional Hospital, Fund for the advancement of health research in Central Denmark Region, the development and resource funds under Danish Regions (*Udviklings- og forskningspuljen ved Danske Regioner*), the Danish Health Confederation (*Sundhedskartellet*), Public Health in the Central Denmark Region – a collaboration between municipalities and the region (*Folkesundhed i Midten*), and Circular Co-Creation under the MedTec Innovation Consortium.

### Competing interests

None declared.

### Patient and public involvement

Patients and the public were not involved in the development of this research cohort.

### Patient consent

Not required.

### Ethical approval

Register-based studies require no ethical approval in Denmark. The study was approved by the Danish Data Protection Agency (file numbers 2007-58-0010 and 2012-58-006) and the Danish Patient Safety Authority (file number 3-3013-1380/1, 3-3013-1380/2, and 3-3013-2741/1).

### Provenance and peer review

Not commissioned; externally peer reviewed.

### Data sharing statement

See Collaboration section.

## Acknowledgements

We gratefully thank the CROSS-TRACKS steering committee for initiating the project and providing access to the cohort. We also thank the Enversion A/S team for data acquisition, modelling, validation, and support.

## Supplementary material

Table 1. Codes for defining population characteristics and healthcare utilisation

Table 2. Cleaning of body mass index registrations from the electronic health record

Table 3. Cleaning of blood pressure registrations from the electronic health record

## Notes

### Competing Interest Statement

The authors have declared no competing interest.

